# Ultrasound Time-Harmonic Elastography: Habitat Viscosity and Tumor Stiffness Heterogeneity for Differentiation of Benign and Malignant Liver Lesions

**DOI:** 10.64898/2026.03.12.26348218

**Authors:** Paul Spiesecke, Mareike Wolff, Thomas Fischer, Ingolf Sack, Tom Meyer

**Affiliations:** Charité - Universitätsmedizin Berlin, corporate member of Freie Universität Berlin, Humboldt-Universität zu Berlin, Department of Radiology, Berlin, Germany; Berlin Institute of Health at Charité – Universitätsmedizin Berlin, BIH Biomedical Innovation Academy, BIH Charité Junior Clinician Scientist Program, Charitéplatz 1, 10117 Berlin, Germany; Charité - Universitätsmedizin Berlin, corporate member of Freie Universität Berlin, Humboldt-Universität zu Berlin, Department of Pediatric Oncology and Hematology, Berlin, Germany

**Keywords:** Ultrasound, elastography, time harmonic elastography, viscosity, heterogeneity, tumor

## Abstract

**Background:** Tumor progression is associated with alterations in tissue mechanical properties. Experimental studies in cancer mechanobiology suggest that increased viscosity of the tumor habitat can promote tumor growth, while malignant tumors often exhibit pronounced mechanical heterogeneity with coexisting soft and rigid regions that facilitate cell motility. Elastography enables noninvasive viscoelastic profiling of soft-tissue properties in vivo and may therefore detect tumor malignancy.

**Purpose:** To investigate whether multiparametric external vibration-based ultrasound time-harmonic elastography (THE) can differentiate benign from malignant liver tumors and identify viscoelastic parameters associated with tumor malignancy.

**Materials and Methods:** In this prospective study conducted from January 2025 to March 2026, 94 patients with focal liver lesions underwent THE. Eighty-four patients were included in the final analysis (41 benign, 39 malignant; 45 women; age range 30-87 years). Liver and tumor stiffness (shear wave speed; SWS), viscosity (loss angle; ϕ), and spatial mechanical heterogeneity (spatial standard deviation, SWS-SD) were quantified. Diagnostic performance for differentiating benign and malignant tumors was assessed using the area under the receiver operating characteristic curve (AUC).

**Results:** Tumor heterogeneity and surrounding habitat viscosity provided the most pronounced differentiation between malignant and benign lesions. Malignant tumors demonstrated higher SWS-SD (0.41±0.20 vs. 0.28±0.11 m/s) and increased ϕ (0.76±0.09 vs. 0.71±0.05 rad) with a combined discriminative power of AUC=0.72. These viscoelastic differences were more pronounced in larger tumors of ≥2.5 cm^2^ area (SWS-SD: 0.47±0.19 vs. 0.32±0.11 m/s; ϕ: 0.78±0.10 vs 0.70±0.04 rad) yielding AUC=0.88 while excellent discriminative power of AUC=0.97 for ≥6 cm^2^ tumor area.

**Conclusion:** Elevated viscosity of the tumor habitat combined with increased tumor stiffness-heterogeneity measured by multiparametric THE can differentiate liver malignancies from benign liver lesions. THE may thus provide a rapid, cost-effective approach for viscoelastic profiling of liver tumors in clinical diagnostic imaging.

## Introduction

Focal liver lesions (FLL) are frequently detected in abdominal imaging either as incidental findings or during staging procedures in patients with known or suspected cancer. Benign FLL are observed in approximately 15% of abdominal ultrasound examinations, with focal fatty sparing and simple cysts accounting for a substantial proportion[1]. In contrast, malignant FLL occur more frequently than primary hepatic malignancies. Population-based analyses indicate that approximately 5% of patients with cancer present with synchronous liver metastases at diagnosis[2]. Accurate differentiation between benign and malignant FLL is therefore essential for guiding patient management and therapy. In clinical practice, lesion characterization is commonly performed using contrast-enhanced imaging techniques such as computed tomography (CT), magnetic resonance imaging (MRI), or contrast-enhanced ultrasound (CEUS))[3]. However, imaging biomarkers reflecting the biomechanical properties of tumors and their surrounding microenvironment provide additional valuable information for lesion characterization[4-6] since tumor progression is associated with alterations in tissue mechanical properties.

Experimental studies in cancer mechanobiology have shown that changes in tissue viscoelasticity influence tumor growth, invasion, and cell motility[7]. In particular, increased viscosity of the surrounding microenvironment may promote tumor progression even in the absence of major stiffness changes[8], while malignant tumors often exhibit pronounced mechanical heterogeneity with coexisting soft and stiff regions[7]. This combination of a permissive tumor habitat and heterogeneous tumor mechanics may facilitate cellular rearrangement and invasive behavior and may therefore serve as an imaging biomarker of tumor aggressiveness[5].

Elastography enables noninvasive quantification of tissue mechanical properties in vivo. Magnetic resonance elastography (MRE)[9] is currently considered the reference standard for viscoelasticity imaging of the liver, despite high costs and limited accessibility[4, 10, 11]. Time-harmonic elastography (THE) is a relatively new, cost-effective, elastography modality that emulates MRE by medical ultrasound as it adopts the MRE principle of externally induced continuous vibrations. THE encodes externally induced shear waves and derives quantitative maps of tissue stiffness and viscosity[12-14]. Uniquely in ultrasound elastography, THE enables full-field-of-view mechanical maps similar to MRE, which is critical for tumor imaging[15] because the tumor and its surrounding habitat tissue are covered by a single scan.

The purpose of this study is to investigate whether multiparametric THE can differentiate benign from malignant focal liver lesions by quantifying tumor stiffness-heterogeneity and viscosity of the surrounding tumor habitat.

## Material and Methods

### Study Design and Subjects

This prospective study was approved by the institutional review board (EA4/165/24). Written informed consent was obtained from all participants prior to study inclusion. Patients with focal liver lesions (FLL) who underwent clinical imaging evaluation at our institution between January 2025 and March 2026 were prospectively recruited. Inclusion criteria were the presence of at least one focal liver lesion detected on ultrasound or cross-sectional imaging and a clinically indicated diagnostic workup for lesion characterization. Exclusion criteria included insufficient shear wave coupling during elastography measurements, defined as less than 50% of the liver or less than 80% of the tumor within the field-of-view meeting the quality criteria described below. A total of 94 patients underwent ultrasound time-harmonic elastography (THE). After quality control and application of the predefined exclusion criteria, 80 patients were included in the final analysis. The final cohort consisted of 41 patients with benign lesions and 39 patients with malignant lesions. The demographical information of the included subjects are summarized in Table 1, the recruitment diagram is shown in Figure 1.

**Table 1.** Demographics, tumor area and viscoelasticity parameters for the tumor entities. Focal fatty sparing, focal fatty infiltration and adenoma were grouped into various benign lesions.

|  | Benign |  |  | Malignant |  |
| --- | --- | --- | --- | --- | --- |
|  | FNH | Hemangioma | Various benign | HCC | Metastases |
| N (women) | 4 (4) | 29 (16) | 8 (6) | 5 (1) | 34 (18) |
| Age (years) | 42 ± 8 | 51 ± 15 | 56 ± 18 | 73 ± 8 | 60 ± 14 |
| **Liver parameters** |  |  |  |  |  |
| Liver SWS (m/s) | 1.13 ± 0.11 | 1.43 ± 0.24 | 1.39 ± 0.19 | 1.71 ± 0.41 | 1.51 ± 0.26 |
| Liver SWS-SD (m/s) | 0.28 ± 0.12 | 0.36 ± 0.1 | 0.39 ± 0.12 | 0.49 ± 0.13 | 0.41 ± 0.14 |
| Liver viscosity $\phi$ (rad) | 0.69 ± 0.05 | 0.71 ± 0.06 | 0.72 ± 0.03 | 0.76 ± 0.1 | 0.76 ± 0.09 |
| Quality area liver (%) | 70 ± 19 | 71 ± 18 | 81 ± 13 | 70 ± 26 | 74 ± 18 |
| **Tumor parameters** |  |  |  |  |  |
| Tumor SWS (m/s) | 1.09 ± 0.07 | 1.63 ± 0.1 | 1.43 ± 0.04 | 1.76 ± 0.1 | 1.6 ± 0.08 |
| Tumor SWS-SD (m/s) | 0.22 ± 0.01 | 0.28 ± 0.12 | 0.3 ± 0.09 | 0.46 ± 0.17 | 0.4 ± 0.2 |
| Tumor viscosity $\phi$ (rad) | 0.74 ± 0.06 | 0.81 ± 0.16 | 0.78 ± 0.06 | 0.87 ± 0.11 | 0.83 ± 0.13 |
| Tumor area (cm<sup>2</sup>) | 13.53 ± 13.4 | 3.42 ± 4.11 | 5.9 ± 2.71 | 9.89 ± 6.83 | 10.88 ± 14.34 |
| Quality area tumor (%) | 94 ± 5 | 97 ± 5 | 96 ± 5 | 93 ± 7 | 95 ± 5 |

**Figure 1.**
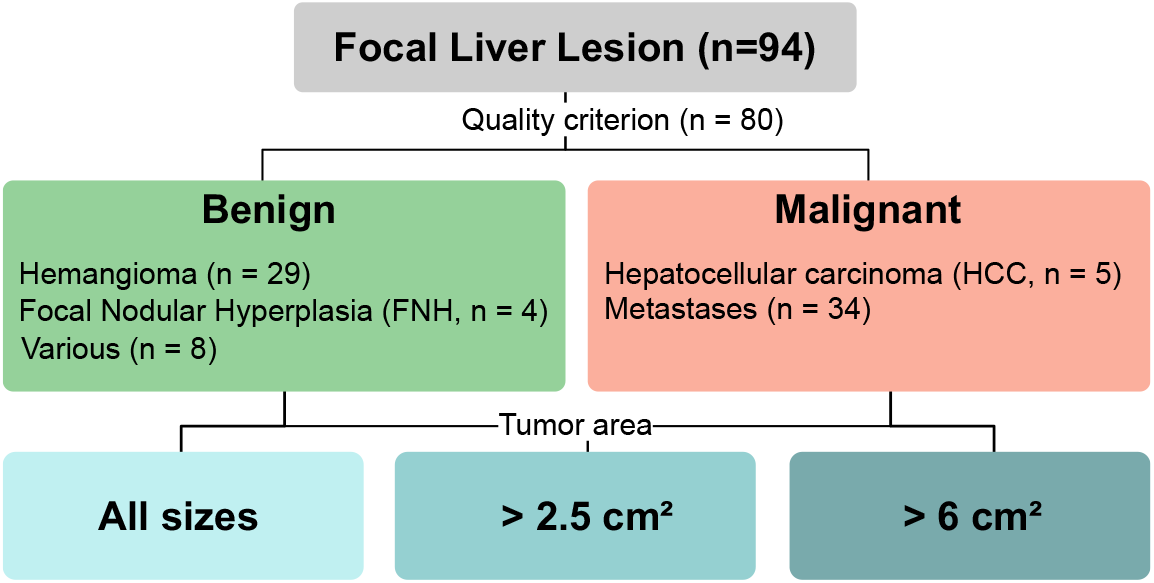
A total of 94 patients with focal liver lesions have been included in the study. The measurements of 14 patients did not meet the quality criteria and were excluded. Focal fatty sparing, focal fatty infiltration and adenoma were grouped into various benign lesions.

### Clinical Workup and Reference Standard

Clinical characterization of focal liver lesions was performed according to institutional diagnostic standards and guidelines. Lesion diagnosis was established using contrast-enhanced ultrasound (CEUS), contrast-enhanced magnetic resonance imaging (MRI), or histopathologic evaluation following core biopsy or surgical resection, depending on the clinical indication. All imaging examinations were reviewed by radiologists with expertise in abdominal imaging. Lesions were classified as benign or malignant based on established imaging criteria or histopathologic findings, respectively. Administration of contrast agents as well as biopsy or surgical procedures were performed solely for clinical indications and were not conducted specifically for study purposes.

### Time-Harmonic Elastography (THE)

To induce harmonic multifrequency vibration (27, 33, 39, 44, 50, and 56 Hz) for time-harmonic elastography (THE), a custom-designed vibration pillow was placed on the standard examination table, and patients were positioned supine on the pillow[14]. The acquisition setup is shown in Figure 2. Ultrasound data were acquired using a clinical ultrasound scanner (GAMPT mbH, Merseburg, Germany) equipped with a 3.3-MHz convex array transducer. Imaging planes were selected to include the focal liver lesion and the surrounding liver parenchyma [12, 13, 16]. Ultrasound radiofrequency (RF) data were recorded over 81 frames at 80 Hz during a 1 s breath hold to capture shear wave propagation. Each acquisition was repeated 10 times.

**Figure 2.**
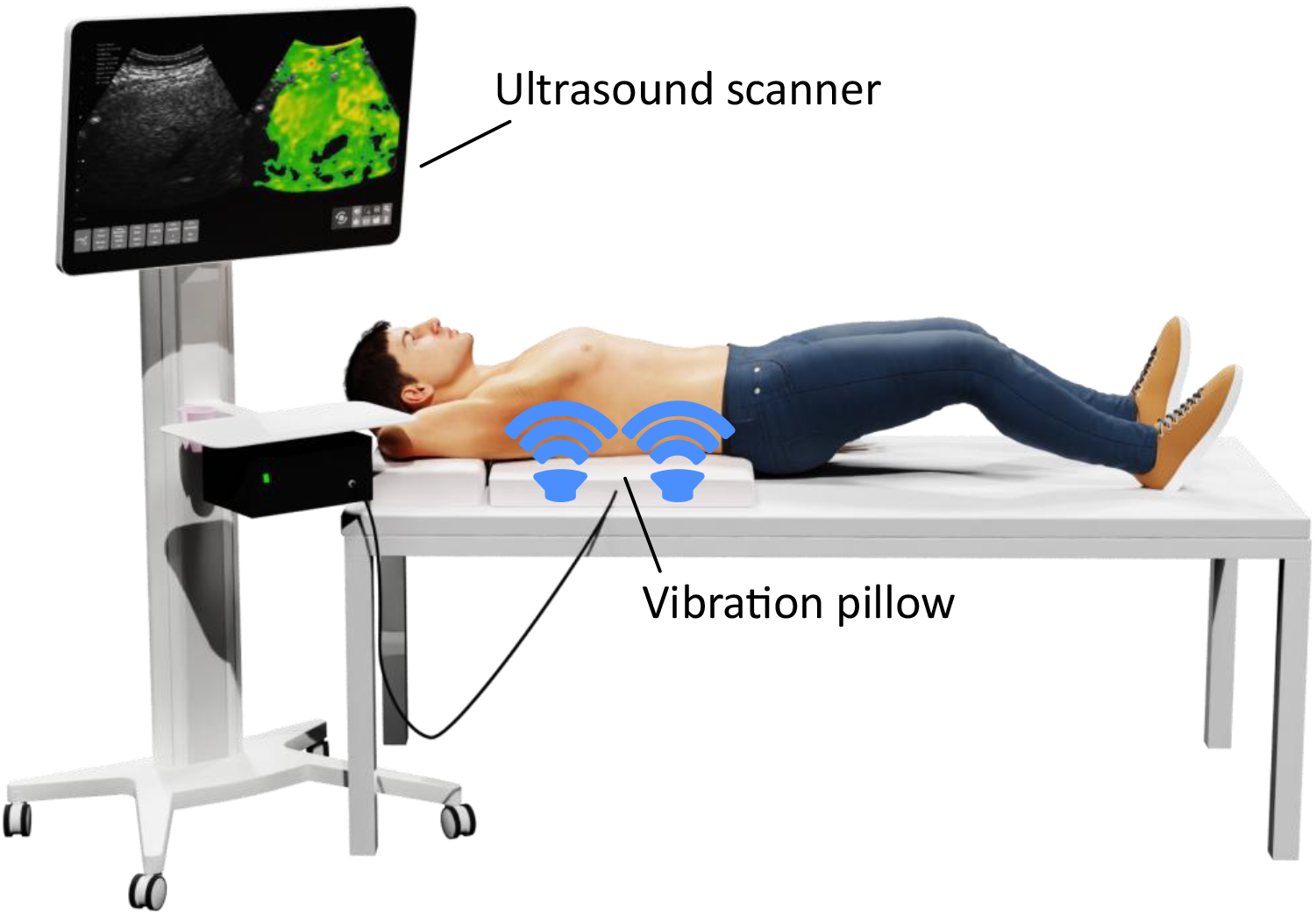
Acquisition setup for external vibration based time harmonic elastography (THE). The setup consists of a clinical ultrasound scanner and a vibration pillow that is placed on the patient bed to induce harmonic multi frequency vibration.

**Figure 3.**
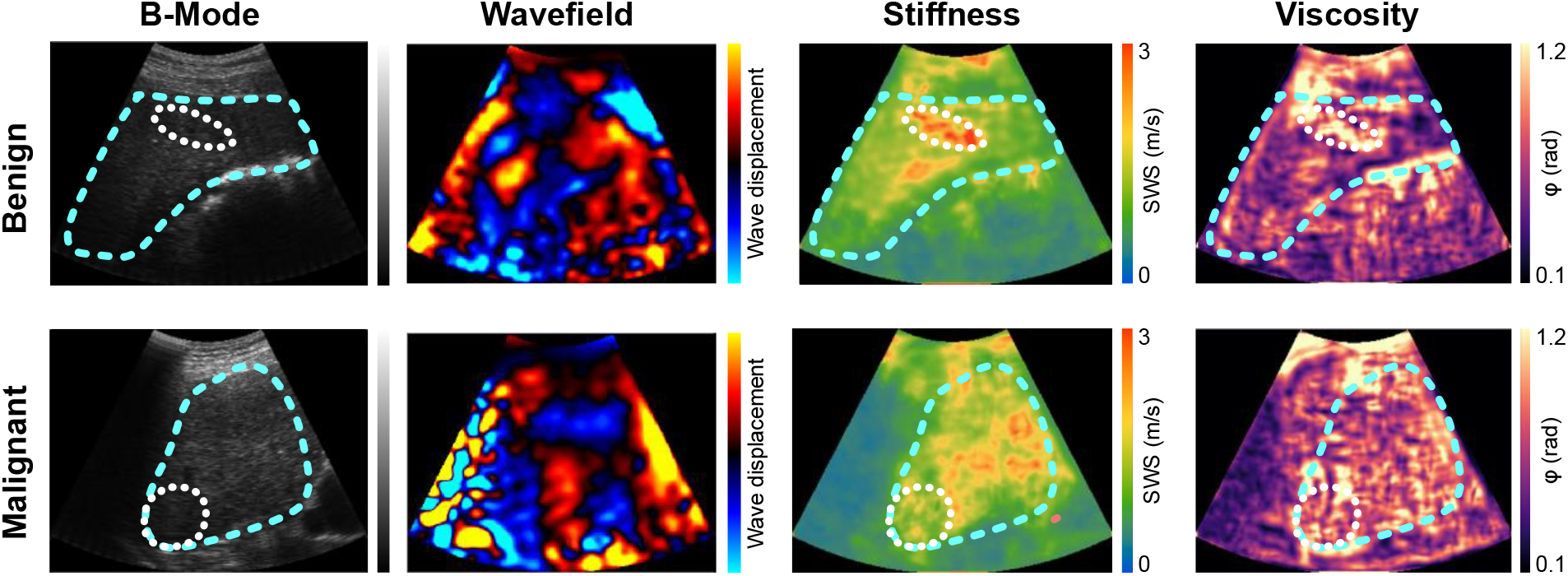
Representative B-mode images, wavefields at 39 Hz vibration, stiffness maps (shear wave speed, SWS), and viscosity maps (loss angle, ϕ) are shown for one subject each with a benign and one subject with a malignant focal liver lesion. The liver region is outlined by a light blue dashed line, and the tumor region by a white dotted line.

Raw ultrasound data were processed using the previously described THE processing pipeline [12, 14]. Briefly, tissue displacement fields were obtained from the RF data using Kasai’s displacement estimation algorithm. A temporal Fourier transform was then applied to isolate harmonic displacement components corresponding to the induced vibration frequencies. Spatially resolved quantitative viscoelastic parameter maps were subsequently reconstructed across the full field of view using wave number (k) based multifrequency gradient inversion (k-MDEV)[17]. This approach yields maps of shear wave speed (SWS) as a surrogate for tissue stiffness and the loss angle of the complex shear modulus (ϕ) as a surrogate for tissue viscosity.

To improve robustness of the inversion, singular value–based denoising was applied to the displacement fields to suppress spatiotemporal noise. Additionally, two-dimensional gradients according to the Andersson method were incorporated into the gradient inversion to further enhance noise stability[18]. Measurement quality was assessed using a composite quality score based on four criteria: (i) correlation coefficient of displacement estimation > 0.9; (ii) spectral z-score > 0.1, reflecting the sharpness of the harmonic peak relative to neighboring frequencies after Fourier transformation; (iii) windowed coefficient of variation of the z-score < 0.5; and (iv) vibration amplitude > 0.5 μm. A frequency-averaged quality score ≥ 3 was considered sufficient for further analysis.

Regions of interest (ROIs) were manually delineated on the corresponding B-mode images using ITK-SNAP (version 3.8). Separate ROIs were defined for the focal liver lesion and the surrounding liver parenchyma representing the tumor habitat.

### Statistical Analysis

Continuous variables are reported as mean ± standard deviation. Differences between benign and malignant lesions were evaluated using t-test. Diagnostic performance for differentiating benign and malignant lesions was assessed using receiver operating characteristic (ROC) analysis, and the area under the curve (AUC) was calculated. A generalized linear model was fitted for the multi parametric classification of tumor malignancy. P < 0.05 was considered statistically significant. All statistical analysis was performed in R (version 4.3.2), using R-Studio (version 2023.12.0, PBC, Boston, MA).

## Results

### Patient Characteristics

Of the 94 patients initially enrolled in the study, 14 examinations were excluded due to insufficient shear wave coupling or failure to meet the predefined elastography quality criteria, described above. Consequently, 80 patients were included in the final analysis. Among them, 39 patients were diagnosed with benign focal liver lesions (FLL) and 41 patients with malignant FLL, according to the reference standard of contrast-enhanced imaging or histopathology. The distribution of lesion entities is summarized in Table 1. Regions not fulfilling the quality criteria were excluded from further analysis by applying the predefined quality mask based on spectral peak sharpness, vibration amplitude, and displacement estimation stability. Mean coverage of liver and tumor regions meeting the quality criteria was 73± 17% and 95 ± 5%% respectively.

### Viscoelasticity Parameters

Representative maps of elastography parameter maps for benign and malignant lesions are shown in Figure 33. All group parameters are summarized in Table 2. Tumor stiffness heterogeneity, expressed as the spatial standard deviation of shear wave speed (SWS-SD), was lower in benign lesions (0.28 ± 0.11 m/s) than in malignant lesions (0.41 ± 0.20 m/s; p <0.05). Both tumor stiffness and viscosity showed no significant difference between the groups. Mean liver viscosity ϕ of the surrounding tumor habitat, was 0.71 ± 0.05 in patients with benign lesions and 0.76 ± 0.09 in patients with malignant lesions. Mean liver stiffness was 1.40 ± 0.14 m/s in benign lesions and 1.53 ± 0.12 m/s in malignant lesions. Overall, tumor stiffness heterogeneity (SWS-SD) and liver viscosity provided the most pronounced differentiation between benign and malignant lesions. Grouped boxplots for malignant and benign lesions are shown in figure 4.

**Table 2.** Demographics and viscoelasticity parameters of malignant and benign lesions obtained by THE and grouped by tumor size. Significant p values are highlighted in bold. The most pronounced group differences for each liver and tumor parameters are underlined.

|  | All tumor sizes |  |  | >2.5 cm<sup>2</sup> |  |  | >6 cm<sup>2</sup> |  |  |
| --- | --- | --- | --- | --- | --- | --- | --- | --- | --- |
|  | Benign | Malignant | p | Benign | Malignant | p | Benign | Malignant | p |
| N (Women) | 41 (26) | 39 (19) |  | 21 (15) | 30 (12) |  | 10 (5) | 18 (9) |  |
| Age (years) | 51 ± 15 | 62 ± 14 | **0.001** | 50 ± 17 | 64 ± 14 | **0.001** | 48 ± 21 | 60 ± 14 | **0.039** |
| **Liver parameters** |  |  |  |  |  |  |  |  |  |
| Liver SWS (m/s) | 1.40 ± 0.24 | 1.53 ± 0.28 | **0.026** | 1.32 ± 0.19 | 1.56 ± 0.30 | **0.001** | 1.31 ± 0.21 | 1.57 ± 0.32 | **0.017** |
| Liver SWS-SD (m/s) | 0.36 ± 0.11 | 0.42 ± 0.14 | **0.028** | 0.33 ± 0.09 | 0.45 ± 0.15 | **0.001** | 0.33 ± 0.1 | 0.44 ± 0.16 | **0.042** |
| Liver viscosity $\phi$ (rad) | 0.71 ± 0.05 | 0.76 ± 0.09 | **0.007** | 0.69 ± 0.04 | 0.78 ± 0.09 | **<0.001** | 0.69 ± 0.04 | 0.79 ± 0.1 | **0.003** |
| Quality area liver (%) | 73 ± 17 | 73 ± 19 | 0.945 | 76 ± 14 | 74 ± 20 | 0.677 | 73 ± 16 | 72 ± 22 | 0.929 |
| **Tumor parameters** |  |  |  |  |  |  |  |  |  |
| Tumor SWS (m/s) | 1.54 ± 0.09 | 1.62 ± 0.08 | 0.311 | 1.45 ± 0.07 | 1.68 ± 0.09 | 0.023 | 1.38 ± 0.07 | 1.61 ± 0.05 | 0.098 |
| Tumor SWS-SD (m/s) | 0.28 ± 0.11 | 0.41 ± 0.2 | **0.001** | 0.32 ± 0.11 | 0.47 ± 0.19 | **0.001** | 0.33 ± 0.12 | 0.48 ± 0.14 | **0.007** |
| Tumor viscosity $\phi$ (rad) | 0.8 ± 0.14 | 0.84 ± 0.13 | 0.195 | 0.8 ± 0.12 | 0.86 ± 0.12 | 0.062 | 0.81 ± 0.14 | 0.87 ± 0.09 | 0.317 |
| Quality area tumor (%) | 96 ± 5 | 94 ± 5 | 0.063 | 95 ± 5 | 93 ± 5 | 0.318 | 92 ± 6 | 94 ± 5 | 0.483 |
| Tumor area (cm<sup>2</sup>) | 5 ± 6 | 11 ± 14 | 0.016 | 8 ± 7 | 14 ± 14 | 0.084 | 13 ± 8 | 20 ± 16 | 0.123 |

**Table 3.**
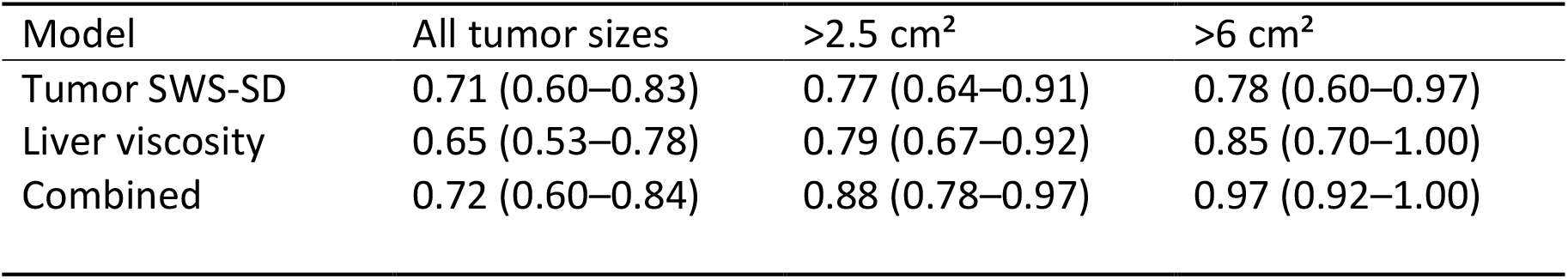
Diagnostic performance quantified as the area under the curve (AUC) of the receiver operating characteristics curve for the discrimination of lesion malignancy for liver viscosity, tumor heterogeneity and a combined model.

| Model | All tumor sizes | >2.5 cm<sup>2</sup> | >6 cm<sup>2</sup> |
| --- | --- | --- | --- |
| Tumor SWS-SD | 0.71 (0.60–0.83) | 0.77 (0.64–0.91) | 0.78 (0.60–0.97) |
| Liver viscosity | 0.65 (0.53–0.78) | 0.79 (0.67–0.92) | 0.85 (0.70–1.00) |
| Combined | 0.72 (0.60–0.84) | 0.88 (0.78–0.97) | 0.97 (0.92–1.00) |

**Figure 4.**
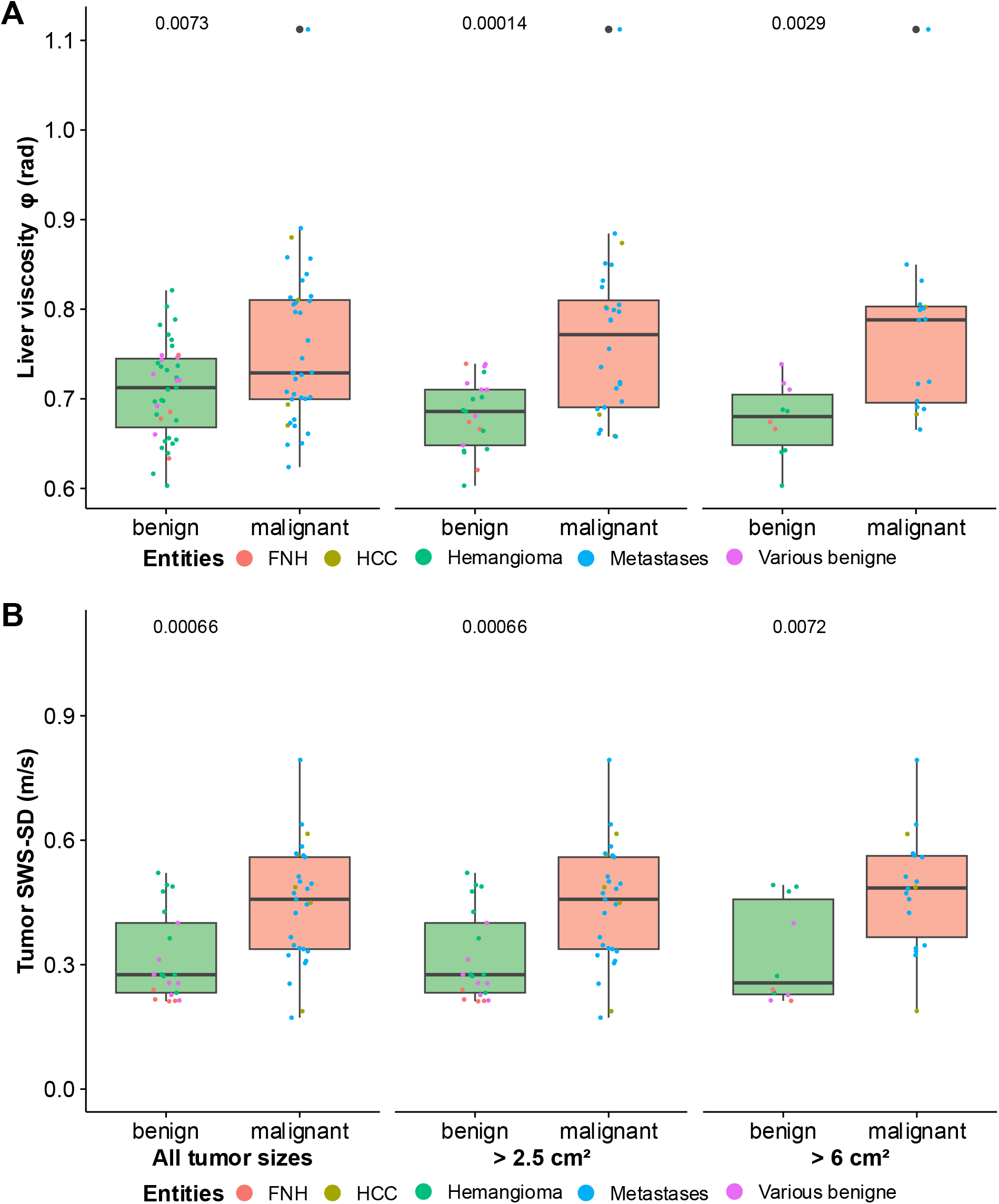
Group comparison of (A) liver viscosity and (B) tumor heterogeneity (SWS-SD) of malignant and benign focal lesions grouped by tumor area.

### Diagnostic Performance

Receiver operating characteristic (ROC) analysis demonstrated good diagnostic accuracy for the discrimination between benign and malignant tumors in a combined linear model for both tumor stiffness heterogeneity and liver tumor habitat viscosity (AUC = 0.72 (95% CI: 0.60-0.84); see figure 5). The diagnostic performance was influenced by tumor size. Restricting the analysis to tumors with a minimal area of ≥ 2.5 cm^2^ resulted in 21 benign and 30 malignant lesions. Under this threshold, elastography parameters showed strengthened group differences (SWS-SD: 0.47 ± 0.19 vs 0.32 ± 0.11, p < 0.001 ϕ: 0.78 ± 0.10 vs 0.69 ± 0.04, p < 0.001) and improved diagnostic performance with an AUC of 0.88 (95% CI: 0.78-0.97). Further restricting the analysis to tumors with a minimal area of ≥ 6 cm^2^ resulted in 10 benign and 18 malignant lesions. Group differences were further strengthened (SWS-SD: 0.48 ± 0.14 vs 0.33 ± 0.12, p < 0.01 ϕ: 0.79 ± 0.10 vs 0.69 ± 0.04, p < 0.01) and diagnostic performance further increased with an AUC of 0.98 (95% CI: 0.92-1.00). The mean tumor size was 5.5 ± 3.0 cm^2^ for benign lesions and 8.0 ± 4.7 cm^2^ for malignant lesions.

**Figure 5.**
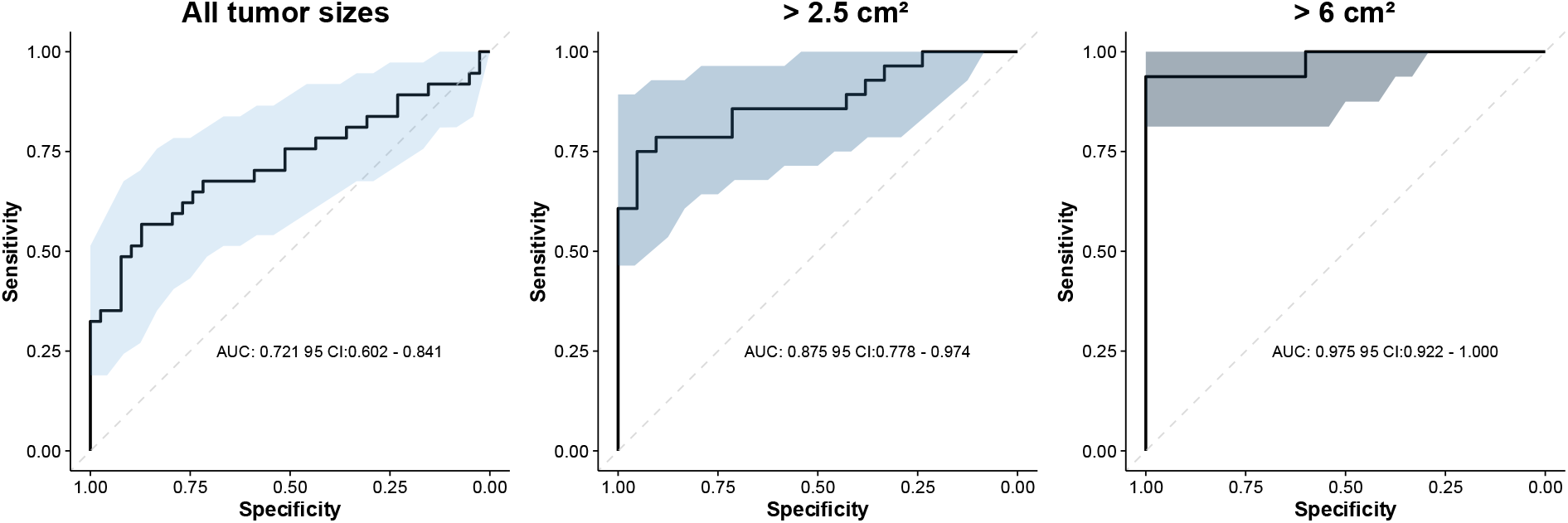
Receiver operating characteristics curves for the differentiation of benign and malignant tumors based on tumor heterogeneity, liver viscosity and a combined prediction of both parameters, grouped by tumor area.

## Discussion

To the best of our knowledge, this is the first study to apply multiparametric THE for the assessment of focal liver lesions in patients. Our results demonstrate that viscosity- and heterogeneity-sensitive THE enables robust quantification of viscoelastic tissue parameters that allow differentiation between benign and malignant liver lesions with good diagnostic accuracy. In particular, tumor stiffness heterogeneity, quantified as the spatial standard deviation of shear wave speed, and the viscosity of the surrounding liver tissue emerged as the most discriminative parameters. This highlights the importance of spatially resolved, viscosity sensitive mapping of tissue mechanical properties with large organ coverage. Diagnostic performance was excellent for larger tumors with an area within the field of view of 6 ≥ cm^2^ and remained good even for smaller lesions with an area between 2.5 and 1 cm^2^. Multiple studies have investigated elastography for the characterization of focal liver lesions[4, 19]. Using ultrasound acoustic radiation force impulse (ARFI)-based shear wave elastography, previous studies reported higher stiffness in malignant liver tumors compared with benign lesions, reflecting increased cellularity and extracellular matrix remodeling[20]. However, the diagnostic performance of ARFI-based ultrasound elastography has been variable across studies, partly due to limited penetration depth and sampling constraints [20-22].

Magnetic resonance elastography (MRE) is considered the reference standard for quantitative viscoelastic imaging of the liver and enables assessment of mechanical tissue properties over large tissue volumes[23]. In particular, Shahryari et al. investigated 77 patients with focal liver lesions using multifrequency MRE and reported good discrimination between benign and malignant tumors based on combined stiffness and viscosity measurements[4]. In viscoelastic materials, the transition from solid-like to fluid-like behavior can be described by the loss angle, which ranges from 0 for purely elastic solids to π/2 for purely viscous fluids. Intermediate values indicate balanced solid–fluid properties, and increasing loss angle reflects greater tissue fluidity, i.e., a stronger viscous contribution to the mechanical response[9]. In that study, malignant lesions showed increased stiffness and higher fluidity compared with surrounding liver tissue, suggesting that solid-fluid interactions within the tumor microenvironment may serve as imaging biomarkers of malignancy. These findings highlight that tumor characterization by elastography should not be limited to bulk stiffness measurements alone but may benefit from parameters reflecting viscoelastic tissue behavior.

Our observations are in line with this concept as we observed increased viscosity in the liver tissue surrounding malignant lesions as well as higher intratumoral stiffness heterogeneity. Elevated habitat viscosity may reflect a more fluid tumor microenvironment that facilitates tumor progression, while increased stiffness heterogeneity may indicate the coexistence of mechanically distinct regions within the tumor architecture. These observations align with recent insights from cancer mechanobiology.[5] Experimental studies have shown that increased tissue fluidity can promote hepatocellular carcinoma growth and invasion even in the absence of major stiffness changes[5, 8]. Furthermore, malignant tumors have been shown to exhibit pronounced mechanical heterogeneity across spatial scales, with coexisting soft and rigid regions that allow tumors to maintain structural integrity while enabling cell motility and invasive behavior[7].

Our study has limitations. First, the spatial resolution of ultrasound elastography imposes constraints on the analysis of very small lesions. As reflected by the improved diagnostic performance observed when restricting the analysis to lesions exceeding a minimal size threshold, small tumors may introduce bias due to partial volume effects and reduced measurement stability. Second, the study cohort included a heterogeneous mix of benign and malignant lesion entities, which may influence the distribution of elastography parameters. Larger studies focusing on specific tumor types will be necessary to further evaluate entity-specific biomechanical signatures.

In summary, the presented study demonstrates the feasibility of multiparametric ultrasound time-harmonic elastography for the biomechanical characterization of focal liver lesions. In particular, increased tumor stiffness heterogeneity and elevated viscosity of the surrounding tumor habitat were associated with malignant lesions and enabled differentiation between benign and malignant tumors with good diagnostic accuracy. These findings support the concept that viscoelastic properties of both the tumor and its microenvironment may serve as imaging biomarkers of tumor malignancy. Given its rapid acquisition, broad availability, and cost-effective implementation using conventional ultrasound systems, time-harmonic elastography may provide a practical approach for biomechanical characterization and differentiation of liver tumors in clinical practice.

## Data Availability

All data produced in the present study are available upon reasonable request to the authors

## Acknowledgements

Funding of the German Research Foundation is gratefully acknowledged (GRK 2260 “BIOQIC”, SFB 1340 “Matrix in Vision”, FOR 5628 “Physics in Cancer”). Dr. Spiesecke is participant in the BIH Charité Junior Clinician Scientist Program funded by the Charité – Universitätsmedizin Berlin, and the Berlin Institute of Health at Charité (BIH).

## Notes

<u>Conflict of interest</u> Thomas Fischer reports having received consultancy honoraria from Bracco and Canon Medical Imaging.

### Competing Interest Statement

Thomas Fischer reports having received consultancy honoraria from Bracco and Canon Medical Imaging.

### Author Declarations

The ethics committe of the Charite - Universitaetsmedizin Berlin gave ethical approval of this work (EA4/165/24).

